# The Dietary Approaches to Stop Hypertension (DASH) diet score and its association with the Risk of Kidney Function Decline and Mortality among Veterans in the Million Veteran Program

**DOI:** 10.64898/2026.08.11.26360178

**Authors:** Jade E. Bowers, Zhihong Yu, Jefferson L. Triozzi, Andrew S. Terker, T Alp Ikizler, Otis Wilson, Kelly Cho, J Michael Gaziano, Ayush Giri, Luis M. Perez, Ran Tao, Christianne L. Roumie, Kerry L. Ivey, Adriana M. Hung

## Abstract

**Background:** The dietary approaches to stop hypertension (DASH) diet is often recommended to patients with chronic kidney disease, although evidence regarding its efficacy in this population is limited. Our study tested the hypothesis that increased adherence to the dietary approaches to stop hypertension (DASH) diet score would be associated with longer time to kidney function decline among Veterans.

**Methods:** We conducted a retrospective cohort study of 251,921 Veterans enrolled in the Million Veteran Program (MVP). The DASH diet score was calculated from the food frequency questionnaire and categorized into tertiles. The primary outcome was a composite of: Kidney event or death, where a kidney event was defined as a sustained 40% decline in estimated glomerular filtration rate (eGFR) or end-stage kidney disease (ESKD). Cox regression models compared the hazard for both outcomes by DASH score tertiles. We examined modification by ancestry, sex and other clinical characteristics

**Results:** The median age was 67 years and 90% of Veterans were men. There were 59,269 (23.5%) who experienced the primary composite outcome, during the maximum follow-up of 10 years (median 6.1 years). Crude incidence rates for the kidney event and death outcome were 43.3, 40.4, and 36.8 per 1000 person-years of DASH score by tertiles. DASH score was associated with a lower hazard ratio (HR) for the primary composite outcome; third vs first tertile 0.81 (95% Confidence Interval (CI) 0.80 – 0.83) and second vs first tertile HR 0.90 [95% CI 0.88 – 0.92]. In subgroup analysis for individuals of African ancestry, Admixed American, and Females, only the third tertile of the DASH score was associated with a statistically significant reduction in composite outcome.

**Conclusion:** Beneficial associations of the DASH diet were observed across subgroups. Future research is needed to understand gene and environmental factors that influence the observed subgroup differences.

## Introduction

Chronic kidney disease (CKD) is a critical global public health issue that impacts approximately 700 million individuals worldwide.^1,2^ There are health disparities involved in the progression of CKD to end-stage kidney disease (ESKD) within individuals of African ancestry who experience a 4-fold higher risk of ESKD.^3^ The impact of CKD compromises a patients’ quality of life and promotes premature death due to cardiovascular complications.^4^

The two most common causes of CKD are diabetes and hypertension.^5^ Dietary modifications are essential for the prevention and treatment of hypertension, obesity, prediabetes, and diabetes, which influence the onset and progression of kidney disease.^6,7,8,9^ ^10^ However, many dietary components have either not been explored or have been sub-optimally explored (magnesium, manganese, etc.) in their relationship to progression of kidney disease.^11,12^ Additionally, it is unknown if the association of dietary factors in kidney disease development or progression varies across populations or if there is effect modification by genes that are involved in CKD, such as the Apolipoprotein L1 gene variant which is associated with an increased risk of kidney disease among individuals of African Ancestry.^13^

The Dietary Approaches to Stop Hypertension (DASH) diet, is a low-sodium diet achieved through increased intake of fruits, vegetables, whole grains, and low-fat dairy products, and used as a treatment adjunct for hypertension in the general population.^14^ DASH diet was originally developed as a dietary intervention to reduce blood pressure, emphasizing limiting sodium intake and increasing the intake of vegetables and whole grains.^15^ Adherence to DASH has since been associated with reduced cardiovascular disease, diabetes in observational studies. Previous studies have suggested an association with decreased chronic kidney disease, although evidence is limited.^15^ The intake profiles of individuals adhering to the DASH diet closely align with the current dietary recommendations for CKD management, such as limiting sodium intake and increasing the intake of vegetables and whole grains.^16^ Despite these potential benefits, research exploring the association between DASH diet adherence and kidney function decline, specifically among Veterans, remains limited. We tested the hypothesis that higher DASH diet score would be associated with lower risk of kidney function decline among veterans in the Million Veteran Program. We also evaluated subgroups by ancestry, age, sex, pre-existing CKD, hypertension, diabetes, and obesity to provide real-world evidence and inform generalizability to clinical practice.

## Methods

### Study Population and Data Sources

This cohort included veterans 18 years or older enrolled in the Million Veteran Program (MVP). The MVP is a national research program designed to examine how genes, lifestyle, military experiences, and environmental factors affect health and wellness in Veterans.^17,18^ MVP enrollment began in early 2011 and remains ongoing. MVP enrolls individuals receiving routine care in Veterans Healthcare Administration and collects data from electronic health records and self-reported MVP surveys. The overall MVP study design has been published, and the details can be found elsewhere.^18^

The MVP Lifestyle Survey includes a semiquantitative food frequency questionnaire (SFFQ).^19^ The date of the completion of the SFFQ was the index date and start of follow-up for those who met all inclusion criteria. We excluded those who did not have a completed MVP Lifestyle Survey or missing answers to questions used for DASH diet score calculation, those with known End Stage Kidney Disease (ESKD) or a renal transplant prior to the survey, missing baseline creatinine, or a baseline estimated glomerular filtration rate (eGFR) less than 30ml/min, those who had records with date errors, or without a last known vital status date after survey.

### Exposure

The exposure was the DASH diet score. Epidemiological scoring methods to quantify DASH diet adherence have been previously validated^20,21^, and subsequently a modified DASH score was developed for use with the MVP lifestyle survey SFFQ and validated in MVP population.^22^ The SFFQ includes 61 food items and assesses the consumption frequency by allowing the participants to choose one of the pre-specified responses: “Never or less than once per month”; “1–3 per month”; “once a week”; “2–4 per week”; “5–6 per week”; “once a day”; “2–3 per day”; “4–5 per day”; and “≥6 per day”. Following the previously published process,^21,22,20^ the modified DASH score was calculated based on 32 questions related to the following seven components: fruits, vegetables, nuts and legumes, low fat dairy products, whole grain, sweetened beverages, and red and processed meats.

### Outcomes

The primary outcome was a composite outcome that included a confirmed 40% decline in eGFR from baseline, ESKD, or mortality. Which is an accepted surrogate endpoint for kidney disease progression as validated by the FDA and adopted in most CKD clinical trials.^23^ The first secondary outcome was restricted to a 40% decline in eGFR or ESKD, and the second secondary outcome was restricted to mortality only. Each eGFR was calculated from serum creatinine using the race-free Chronic Kidney Disease Epidemiology Collaboration equation.^24^ Baseline creatinine was obtained as the closest measure before the SFFQ index date and up to 730 days prior to the index date. To determine the eGFR decline of 40% we utilized only outpatient creatinine measurements from the clinical laboratory results in the EHR. We restricted to one creatinine per day during the follow-up time frame and used the following rules: (1) If a participant had multiple measurements on a given day and the difference between the trough and peak of creatinine measurements was ≤0.3 ml/dl, the average of creatinine measurements for that day was retained for eGFR estimation, and (2) if the difference was >0.3 ml/dl, measurements from that day were excluded. Two eGFR measure were observed during the follow-up period to assess if there had been any decline. A sustained eGFR decline of 40% had to be confirmed by a second eGFR between 3 and 12 months later. The first confirmed event was the event date. This method avoids capturing a single reduced eGFR which may represent acute kidney injury.

ESKD was defined as reaching one of the following: an eGFR of 15 ml/min per 1.73 m^2^, or the first inpatient or outpatient code for dialysis, or related procedures on two different occasions more than 90 days apart, or renal transplantation, which did not require a confirmatory code. (Supplemental Table 1)

### Covariates

Characteristics were collected at the time of the SFFQ completion and included demographics (age, sex, and genetic ancestry), clinical variables (creatinine, eGFR, Body Mass Index (BMI), systolic and diastolic blood pressure, albuminuria, and proteinuria from dipstick), comorbidities (diabetes, hypertension, congestive heart failure, cardiovascular disease, CKD, and kidney stone), and social determinants of health (homelessness, income, alcohol consumption, smoking status, married status). The information on all covariates was collected from the measurement closest to and preceding the MVP Lifestyle Survey.

Data on covariates was extracted from the EHR records,^25^ Census Data, MVP study mart and MVP Baseline and MVP Lifestyle Surveys.^17^ Comorbidities were defined using the *International Classification of Diseases, Ninth Revision, Tenth Revision (ICD-9, ICD-10)*, procedures codes ICD-9/ICD-10 and Current Procedural Terminology (CPT) codes. Covariate definitions are included in Supplemental Table 2. Ancestry was genetically inferred using the 30 top principal ancestry components, and ancestry markers were derived from the 1000 genome project.^26^

### Statistical Analysis

Cox proportional hazard models were used to study the association between the DASH diet score on the primary and secondary outcome. The DASH score was categorized into tertiles and the first tertile (lowest DASH diet score) was used as the referent group. Nested multivariable models were sequentially constructed (a) model 1, minimally adjusted (age, sex, and ancestry), (b) model 2, added baseline eGFR, BMI, and comorbidities (pre-existing CKD, diabetes, and hypertension), (c) model 3, additionally adjusted for social determinants of health including income, marital status, alcohol consumption frequency, smoking, and homeless status. The natural spline with 4 degrees of freedom was applied to all continuous variables. The proportional hazards assumption was verified graphically using Schoenfeld residual plots

In consideration of the potential differential associations of DASH diet on kidney disease progression for specific subgroups, we performed *a priori* defined subgroup analysis defined by ancestry, sex, pre-existing CKD, hypertension, diabetes, obesity (BMI < 25, 25-30, and >30 kg/m²), and age (< 65, 65-75 and >75 years). Statistical significance of the interaction term of DASH with each subgroup variable was tested. All statistical tests were 2-sided, where a P < 0.05 or a 95% CI that did not contain unity were considered statistically significant. All analyses were conducted using R statistical software (version 4.4.1.; R Foundation, Inc).^27^

## Results

### Analytic Cohort and Cohort characteristics

In total, 447,259 veterans completed the SFFQ by the end of January 2023, and 251,921 were included in the analytic cohort after applying exclusion criteria (**Figure 1**). Among the 251,921 participants, the median age was 67 years with an interquartile range (IQR) of 59 to 72 years. Veterans were 61.8% European descent and 7.9% African ancestry. The majority of the population (90.0%) was male (**Table 1**). On the index date, Veterans commonly had a diagnosis of diabetes (27.5%), hypertension (63.5%), and cardiovascular disease (72.4%). Veterans were often in the $50,000 or less income bracket (44.7%), stated that they were married (51.8%), never consume alcohol (32.9%), and were former smokers (41.7%). There were no clinically significant differences in Veteran characteristics by DASH diet score tertile.

**Figure 1.**
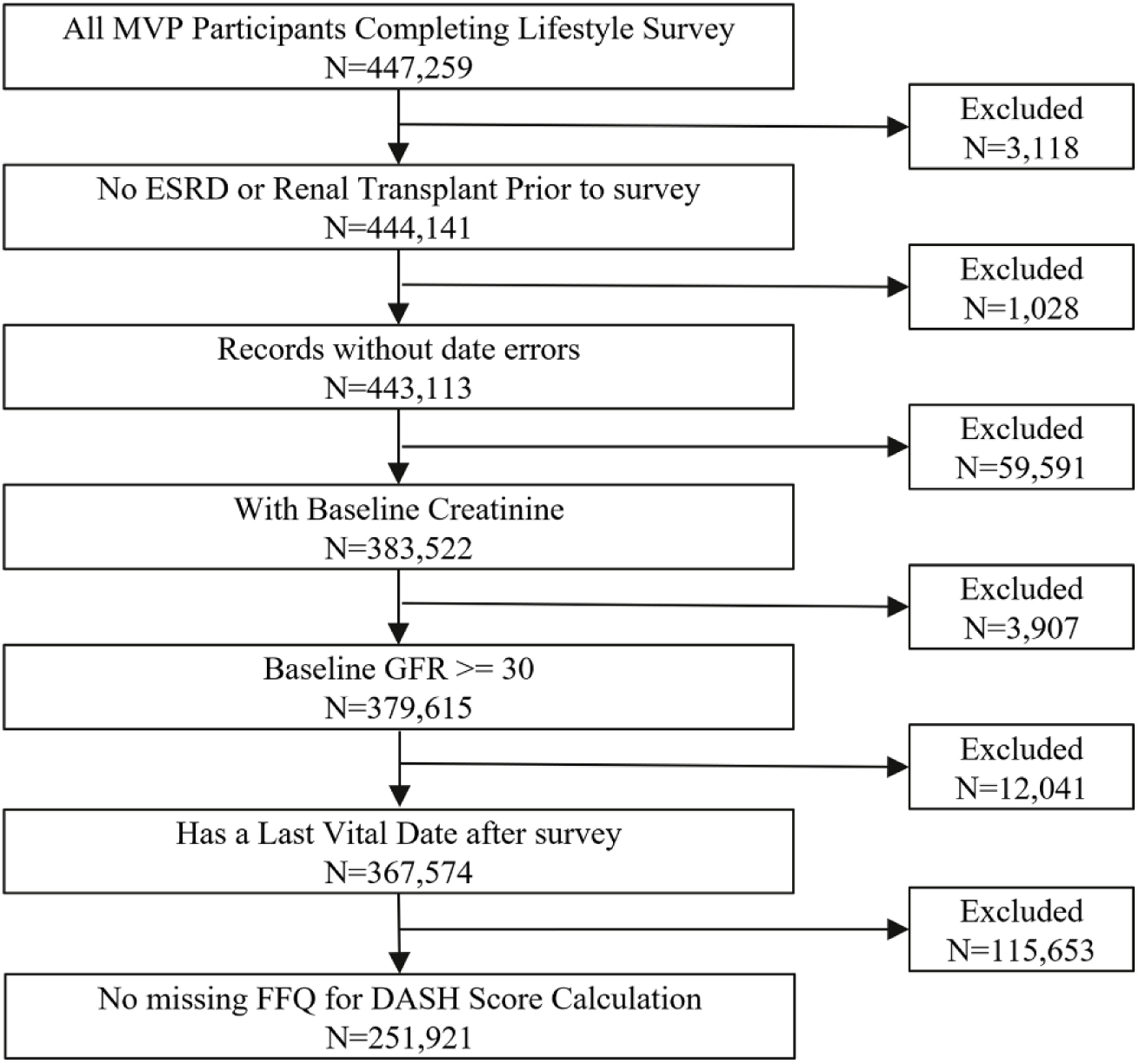
Flowchart for generating the study cohort.

**Table 1:**
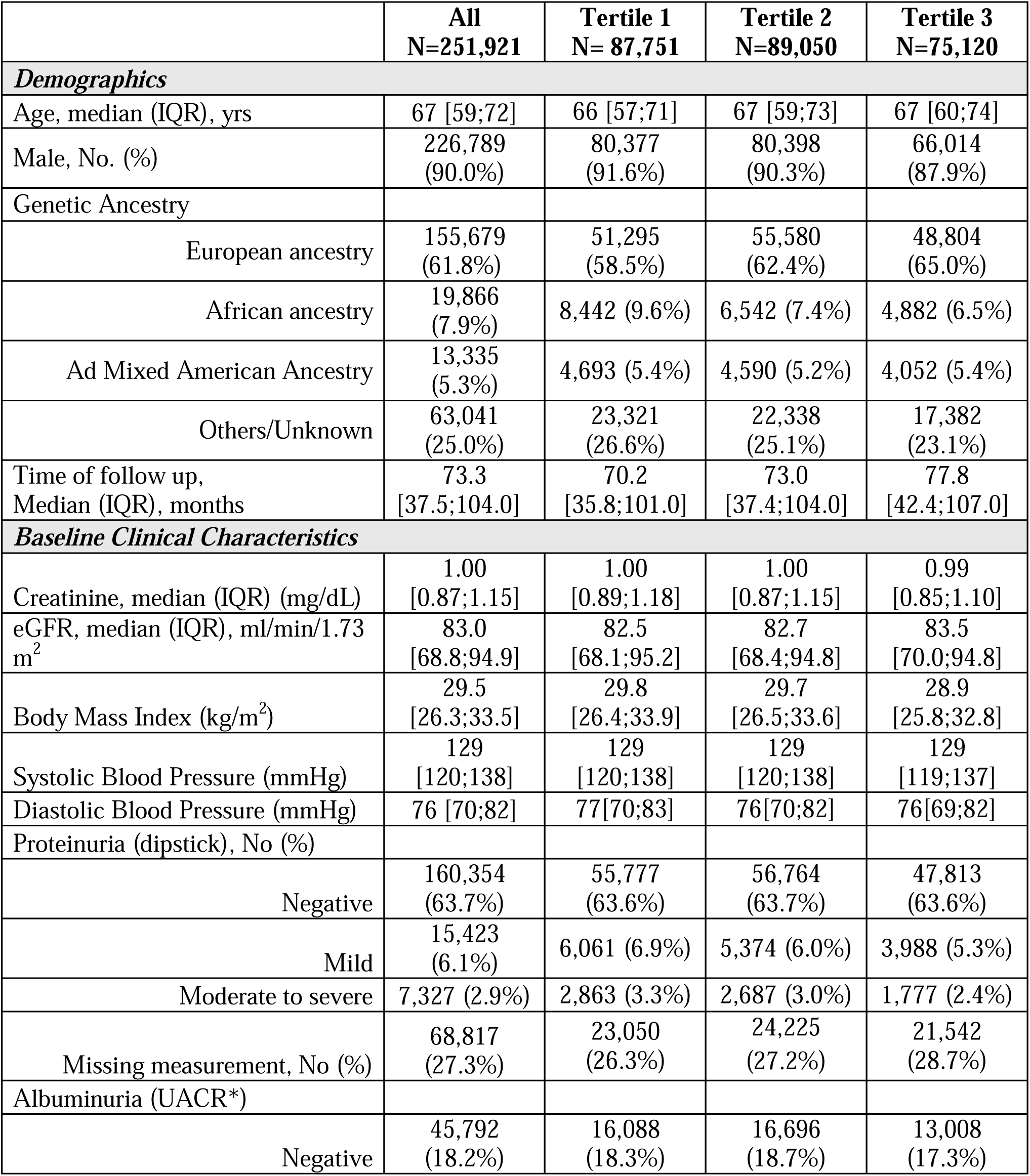
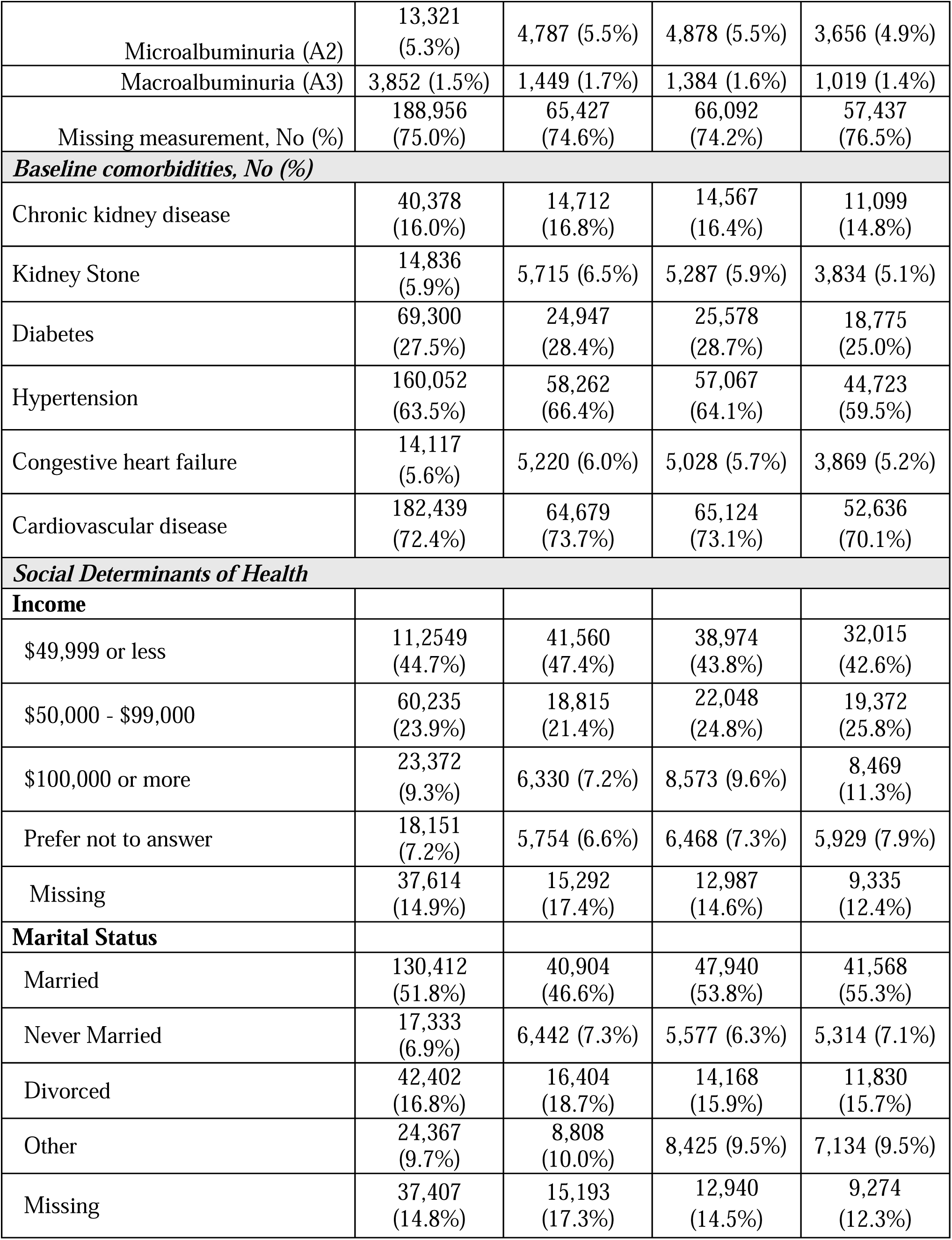
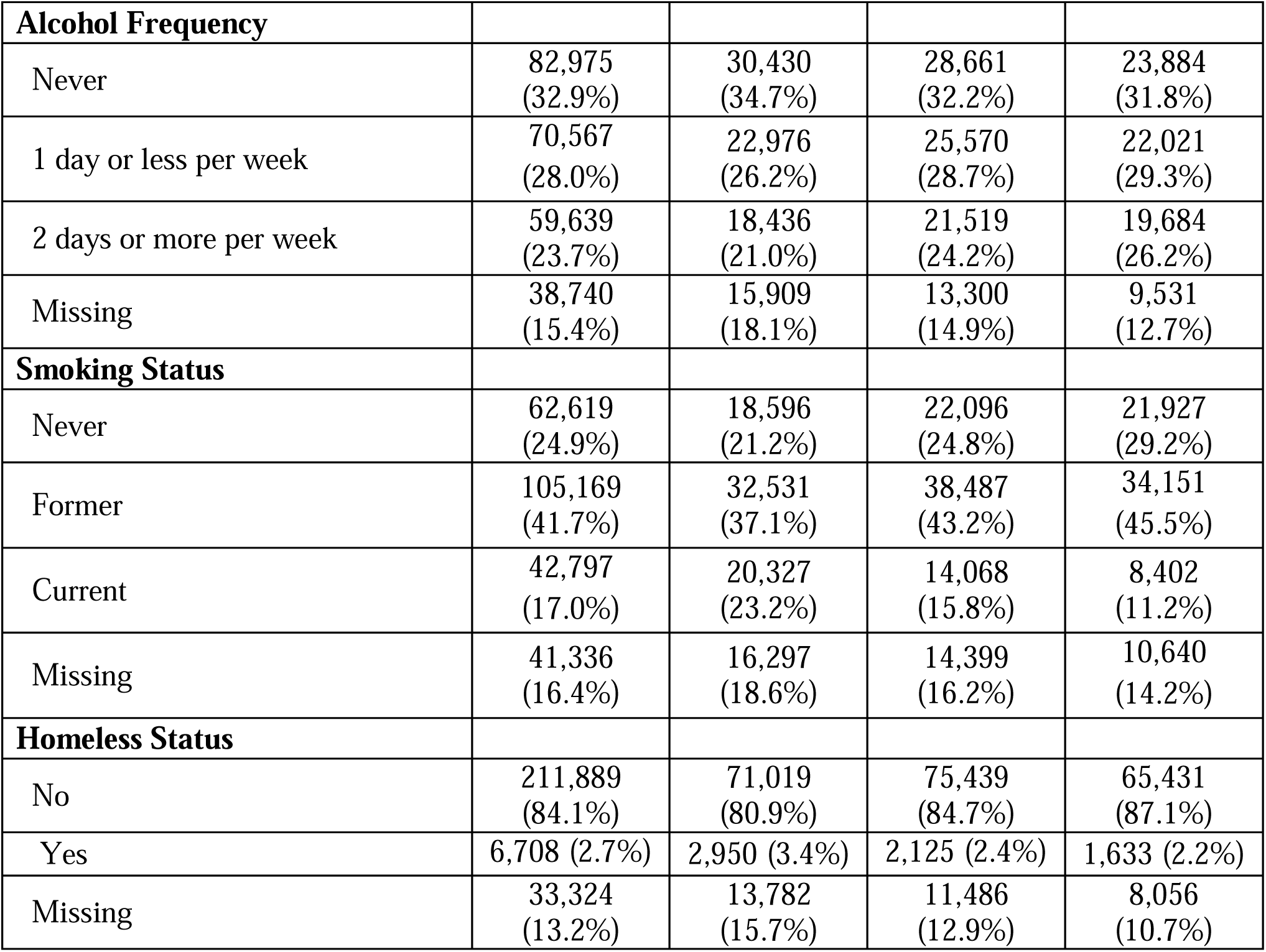
Patient Characteristics at the time of completion of the Food Frequency Questionnaire.

|  | **All**<br>**N=251,921** | **Tertile 1**<br>**N= 87,751** | **Tertile 2**<br>**N=89,050** | **Tertile 3**<br>**N=75,120** |
| --- | --- | --- | --- | --- |
| **Demographics** |  |  |  |  |
| Age, median (IQR), yrs | 67 [59;72] | 66 [57;71] | 67 [59;73] | 67 [60;74] |
| Male, No. (%) | 226,789<br>(90.0%) | 80,377<br>(91.6%) | 80,398<br>(90.3%) | 66,014<br>(87.9%) |
| Genetic Ancestry |  |  |  |  |
| European ancestry | 155,679<br>(61.8%) | 51,295<br>(58.5%) | 55,580<br>(62.4%) | 48,804<br>(65.0%) |
| African ancestry | 19,866<br>(7.9%) | 8,442 (9.6%) | 6,542 (7.4%) | 4,882 (6.5%) |
| Ad Mixed American Ancestry | 13,335<br>(5.3%) | 4,693 (5.4%) | 4,590 (5.2%) | 4,052 (5.4%) |
| Others/Unknown | 63,041<br>(25.0%) | 23,321<br>(26.6%) | 22,338<br>(25.1%) | 17,382<br>(23.1%) |
| Time of follow up,<br>Median (IQR), months | 73.3<br>[37.5;104.0] | 70.2<br>[35.8;101.0] | 73.0<br>[37.4;104.0] | 77.8<br>[42.4;107.0] |
| **Baseline Clinical Characteristics** |  |  |  |  |
| Creatinine, median (IQR) (mg/dL) | 1.00<br>[0.87;1.15] | 1.00<br>[0.89;1.18] | 1.00<br>[0.87;1.15] | 0.99<br>[0.85;1.10] |
| eGFR, median (IQR), ml/min/1.73<br>m<sup>2</sup> | 83.0<br>[68.8;94.9] | 82.5<br>[68.1;95.2] | 82.7<br>[68.4;94.8] | 83.5<br>[70.0;94.8] |
| Body Mass Index (kg/m<sup>2</sup>) | 29.5<br>[26.3;33.5] | 29.8<br>[26.4;33.9] | 29.7<br>[26.5;33.6] | 28.9<br>[25.8;32.8] |
| Systolic Blood Pressure (mmHg) | 129<br>[120;138] | 129<br>[120;138] | 129<br>[120;138] | 129<br>[119;137] |
| Diastolic Blood Pressure (mmHg) | 76 [70;82] | 77[70;83] | 76[70;82] | 76[69;82] |
| Proteinuria (dipstick), No (%) |  |  |  |  |
| Negative | 160,354<br>(63.7%) | 55,777<br>(63.6%) | 56,764<br>(63.7%) | 47,813<br>(63.6%) |
| Mild | 15,423<br>(6.1%) | 6,061 (6.9%) | 5,374 (6.0%) | 3,988 (5.3%) |
| Moderate to severe | 7,327 (2.9%) | 2,863 (3.3%) | 2,687 (3.0%) | 1,777 (2.4%) |
| Missing measurement, No (%) | 68,817<br>(27.3%) | 23,050<br>(26.3%) | 24,225<br>(27.2%) | 21,542<br>(28.7%) |
| Albuminuria (UACR*) |  |  |  |  |
| Negative | 45,792<br>(18.2%) | 16,088<br>(18.3%) | 16,696<br>(18.7%) | 13,008<br>(17.3%) |
| Microalbuminuria (A2) | 13,321<br>(5.3%) | 4,787 (5.5%) | 4,878 (5.5%) | 3,656 (4.9%) |
| Macroalbuminuria (A3) | 3,852 (1.5%) | 1,449 (1.7%) | 1,384 (1.6%) | 1,019 (1.4%) |
| Missing measurement, No (%) | 188,956<br>(75.0%) | 65,427<br>(74.6%) | 66,092<br>(74.2%) | 57,437<br>(76.5%) |
| **Baseline comorbidities, No (%)** |  |  |  |  |
| Chronic kidney disease | 40,378<br>(16.0%) | 14,712<br>(16.8%) | 14,567<br>(16.4%) | 11,099<br>(14.8%) |
| Kidney Stone | 14,836<br>(5.9%) | 5,715 (6.5%) | 5,287 (5.9%) | 3,834 (5.1%) |
| Diabetes | 69,300<br>(27.5%) | 24,947<br>(28.4%) | 25,578<br>(28.7%) | 18,775<br>(25.0%) |
| Hypertension | 160,052<br>(63.5%) | 58,262<br>(66.4%) | 57,067<br>(64.1%) | 44,723<br>(59.5%) |
| Congestive heart failure | 14,117<br>(5.6%) | 5,220 (6.0%) | 5,028 (5.7%) | 3,869 (5.2%) |
| Cardiovascular disease | 182,439<br>(72.4%) | 64,679<br>(73.7%) | 65,124<br>(73.1%) | 52,636<br>(70.1%) |
| **Social Determinants of Health** |  |  |  |  |
| **Income** |  |  |  |  |
| \$49,999 or less | 11,2549<br>(44.7%) | 41,560<br>(47.4%) | 38,974<br>(43.8%) | 32,015<br>(42.6%) |
| \$50,000 - \$99,000 | 60,235<br>(23.9%) | 18,815<br>(21.4%) | 22,048<br>(24.8%) | 19,372<br>(25.8%) |
| \$100,000 or more | 23,372<br>(9.3%) | 6,330 (7.2%) | 8,573 (9.6%) | 8,469<br>(11.3%) |
| Prefer not to answer | 18,151<br>(7.2%) | 5,754 (6.6%) | 6,468 (7.3%) | 5,929 (7.9%) |
| Missing | 37,614<br>(14.9%) | 15,292<br>(17.4%) | 12,987<br>(14.6%) | 9,335<br>(12.4%) |
| **Marital Status** |  |  |  |  |
| Married | 130,412<br>(51.8%) | 40,904<br>(46.6%) | 47,940<br>(53.8%) | 41,568<br>(55.3%) |
| Never Married | 17,333<br>(6.9%) | 6,442 (7.3%) | 5,577 (6.3%) | 5,314 (7.1%) |
| Divorced | 42,402<br>(16.8%) | 16,404<br>(18.7%) | 14,168<br>(15.9%) | 11,830<br>(15.7%) |
| Other | 24,367<br>(9.7%) | 8,808<br>(10.0%) | 8,425 (9.5%) | 7,134 (9.5%) |
| Missing | 37,407<br>(14.8%) | 15,193<br>(17.3%) | 12,940<br>(14.5%) | 9,274<br>(12.3%) |
| **Alcohol Frequency** |  |  |  |  |
| Never | 82,975<br>(32.9%) | 30,430<br>(34.7%) | 28,661<br>(32.2%) | 23,884<br>(31.8%) |
| 1 day or less per week | 70,567<br>(28.0%) | 22,976<br>(26.2%) | 25,570<br>(28.7%) | 22,021<br>(29.3%) |
| 2 days or more per week | 59,639<br>(23.7%) | 18,436<br>(21.0%) | 21,519<br>(24.2%) | 19,684<br>(26.2%) |
| Missing | 38,740<br>(15.4%) | 15,909<br>(18.1%) | 13,300<br>(14.9%) | 9,531<br>(12.7%) |
| **Smoking Status** |  |  |  |  |
| Never | 62,619<br>(24.9%) | 18,596<br>(21.2%) | 22,096<br>(24.8%) | 21,927<br>(29.2%) |
| Former | 105,169<br>(41.7%) | 32,531<br>(37.1%) | 38,487<br>(43.2%) | 34,151<br>(45.5%) |
| Current | 42,797<br>(17.0%) | 20,327<br>(23.2%) | 14,068<br>(15.8%) | 8,402<br>(11.2%) |
| Missing | 41,336<br>(16.4%) | 16,297<br>(18.6%) | 14,399<br>(16.2%) | 10,640<br>(14.2%) |
| **Homeless Status** |  |  |  |  |
| No | 211,889<br>(84.1%) | 71,019<br>(80.9%) | 75,439<br>(84.7%) | 65,431<br>(87.1%) |
| Yes | 6,708 (2.7%) | 2,950 (3.4%) | 2,125 (2.4%) | 1,633 (2.2%) |
| Missing | 33,324<br>(13.2%) | 13,782<br>(15.7%) | 11,486<br>(12.9%) | 8,056<br>(10.7%) |

### Association of DASH diet score with primary and secondary outcome

The crude incidence rates for the primary outcome were 43.3, 40.4, and 36.8 per 1000 person-years for tertiles 1-3 of DASH diet score. Higher DASH scores were statistically significantly associated with a reduction in both the primary outcome (40% eGFR decline or ESKD or mortality) and the secondary outcomes (the composite of 40% eGFR decline or ESKD and mortality separately), in all nested models (**Table 2**). When we analyzed the association between the DASH diet score and the primary outcome, we found that the participants in the second and third tertile had a 9.9% or 18.6% lower association with the primary outcome (HR: 0.901, 95% CI: 0.884 – 0.919 or 0.814, 95% CI: 0.797 – 0.831, respectively) compared to those in the first tertile, after adjusting for the prespecified demographic, clinical factors, and social determinants of health.

**Table 2:**
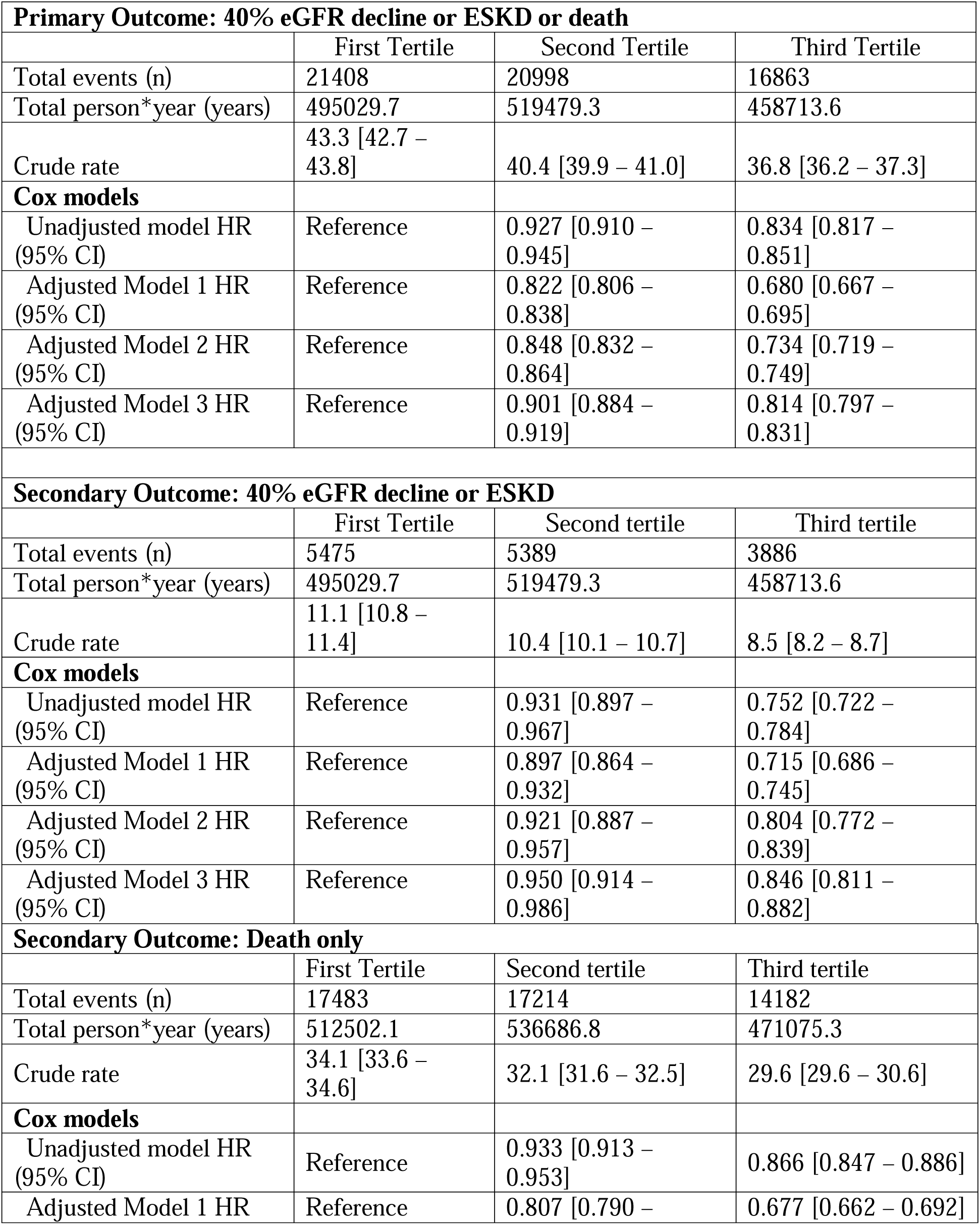
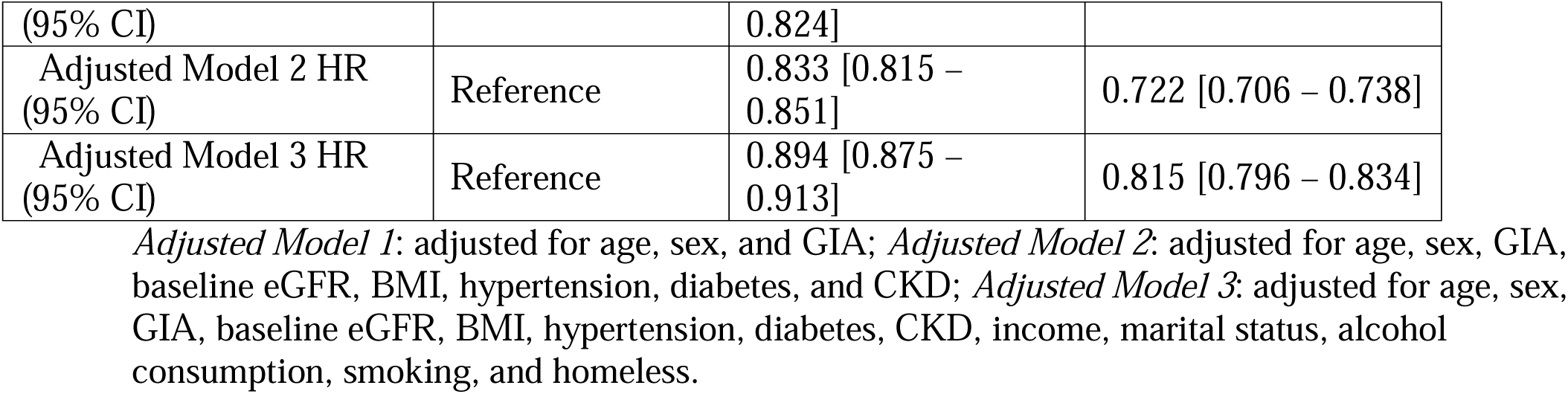
Crude Rates and Hazard Ratio with 95% Confidence Interval for Primary and Secondary Outcomes.

For the first secondary outcome of CKD, the crude incidence rates were 11.1, 10.4, and 8.5, and for the second secondary outcome of death the crude incidence rates were 34.1, 32.1, and 29.6. Similarly, for the primary secondary outcome of kidney events, after adjusting for covariates, Veterans in the second or third tertile of DASH diet score demonstrated a respective 5.0% or 15.4% lower hazard (HR: 0.950, 95% CI: 0.914 – 0.986 or 0.846, 95% CI: 0.811 – 0.882, respectively) compared to those in the first tertile. Additionally, for the second secondary outcome, after adjusting for covariates, individuals in the second or third tertile of DASH diet score portrayed a 10.6% or 18.5% lower hazard (HR: 0.894, 95% CI: 0.875 – 0.913 or 0.815, 95% CI: 0.796 – 0.834, respectively) compared to participants in the first tertile.

The cumulative incidence curves stratified by DASH score tertiles separated by two years and demonstrated a reduction in the cumulative incidence of kidney event and/or death between the first to the third tertile for both primary and secondary outcomes (**Figure 2 panels A -C**).

**Figure 2.**
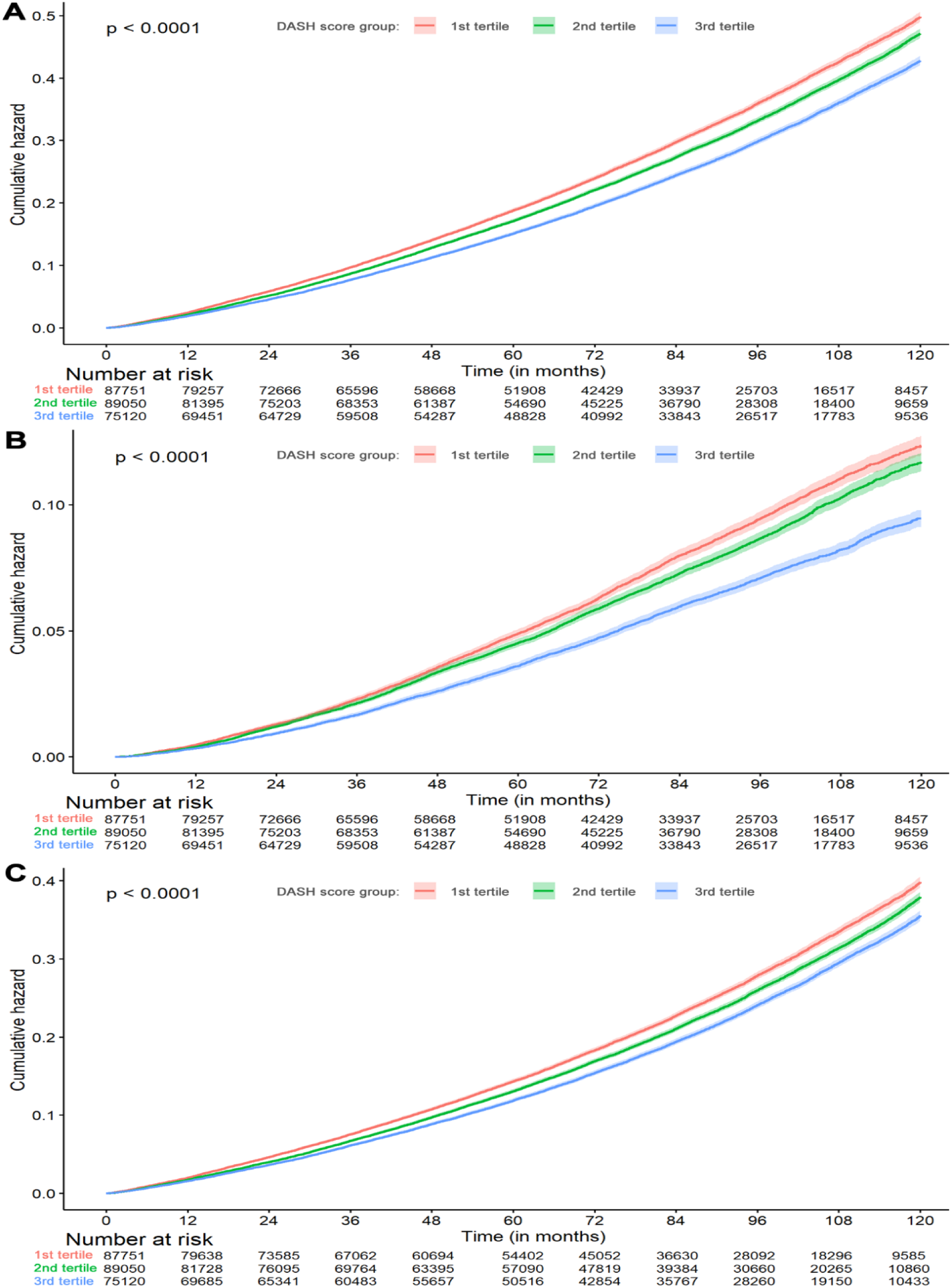
Cumulative hazard by DASH score tertiles for (A): the primary outcome of the kidney composite (i.e., 40% decline in eGFR or ESKD) or death; (B): the secondary outcome of kidney composite; (C): the secondary outcome of death.

### Subgroup analysis

We examined the relationship between the DASH diet score and the primary and secondary outcomes across subgroups. For the primary outcome, DASH diet adherence in the third or second tertile was associated with a significant protective association across most subgroups. Notably, the protective association in the third tertile was significant in all examined subgroups (**Figure 3**).

**Figure 3.**
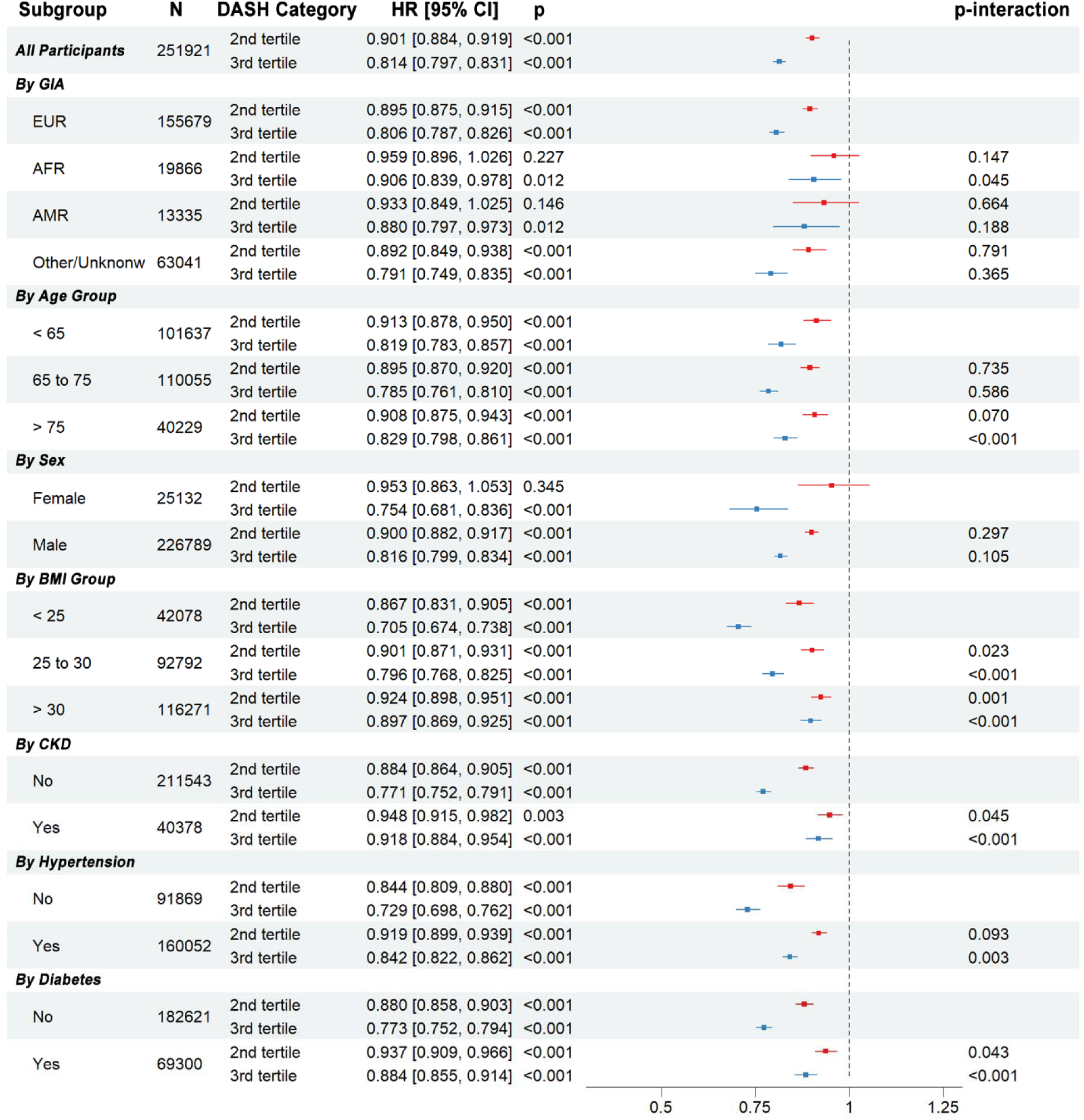
Forest plot presenting the hazard ratio from Adjusted Model 3 for 40% eGFR decline, ESKD or death by DASH score tertiles (1st tertile as reference) among all participants and different subgroups.

Among subgroups by baseline comorbidities, DASH score demonstrated a stronger protective association in participants without diabetes (HR: 0.773, 95% CI: 0.752-0.794) in comparison to those with diabetes (HR: 0.884, 95% CI: 0.855-0.914; p-interaction < 0.001). Similarly, the DASH diet score in participants without hypertension showed a larger protective association (HR: 0.729, 95% CI: 0.698-0.762) compared to those with hypertension (HR: 0.842, 95% CI: 0.822-0.862; p-interaction = 0.003). The DASH diet score in participants without pre-existing CKD also exhibited stronger protective association (HR: 0.771, 95% CI: 0.752-0.791) compared to those with CKD at baseline (HR: 0.918, 95% CI: 0.884-0.954; p-interaction < 0.001). To assess the association by obesity, participants were categorized into three baseline BMI groups, and the strongest protective association of the DASH score was observed in the non-obese group (BMI <25 kg/m2; HR: 0.705, 95% CI: 0.674-0.738) compared to the obese group (BMI >30kg/m2; HR: 0.897, 95% CI: 0.869-0.925; p-interaction < 0.001).

Veterans of European ancestry experienced a statistically significant protective association (HR: 0.806, 95% CI: 0.787-0.826) compared to individuals of African ancestry (HR:0.906, 95% CI: 0.839-0.978, p-interaction = 0.045). The protective association was consistent between female (HR: 0.754, 95% CI: 0.681-0.836) and male participants (HR: 0.816, CI: 0.799-0.834), p-interaction of 0.105.

### Correlation between adherence to the DASH diet score and intake of characterizing nutrients

We conducted a partial correlation analysis to examine the correlation between DASH score and several nutrients including calories, animal protein, sodium, potassium, phosphorus, magnesium, manganese, and calcium (**Figure 4**). We found that higher intakes of potassium, magnesium, and manganese were positively associated with higher adherence to the DASH diet score. This positive correlation might associate the protective impact of DASH score in reducing the kidney disease progression or death. In contrast, higher intake of sodium was negatively correlated with adherence to the DASH diet score, indicating that higher sodium intake was linked to a lower DASH score and consequently may be associated with reduced protective benefits for kidney disease.

**Figure 4.**
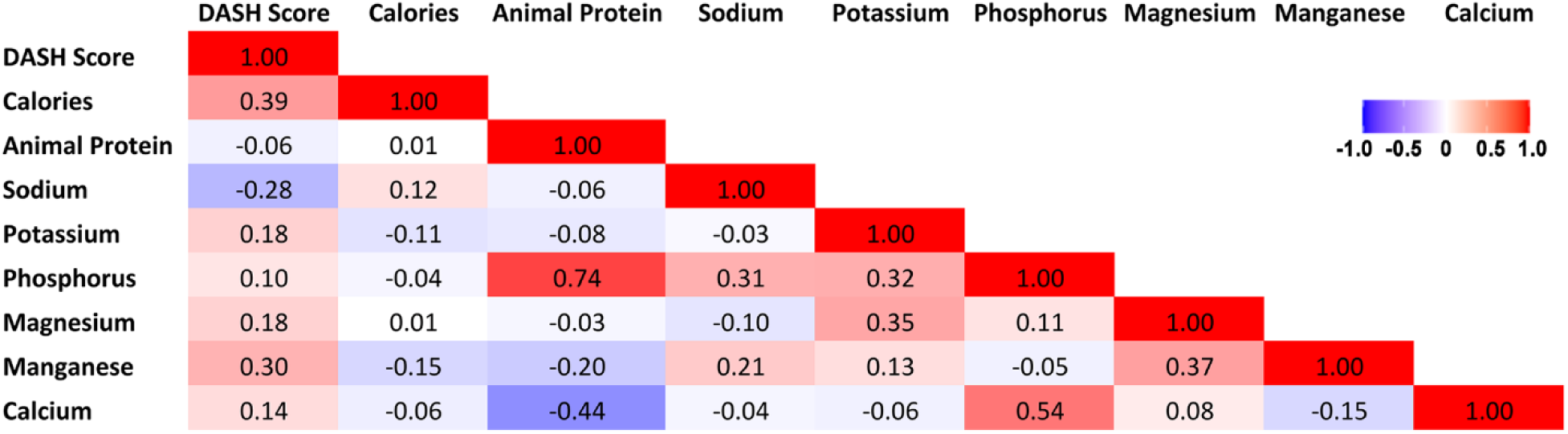
Heatmap of partial correlation coefficients between DASH score and nutrients.

## Discussion

Higher adherence to the DASH dietary pattern was associated with a lower hazard of kidney disease progression, death, and their composite across all adjusted models. Veterans in the highest tertile demonstrated an 18.6% lower hazard for the primary composite outcome compared to those in the lowest tertile, corresponding to an absolute reduction of 6.5 events per 1000 person-years. Stated another way, for every 154 participants that highly adhere to the DASH diet for one year, one event (CKD and death) could be prevented. A consistent pattern was observed in the secondary outcomes. These findings have meaningful public health implications given the disproportionate burden of CKD and its complications among United States Veterans.^28^

The stronger protective association observed among Veterans without pre-existing diabetes, hypertension, or CKD suggests that DASH diet adherence may be most impactful as primary prevention. This is potentially due to previously published literature of the DASH diet’s ability to prevent hypertension and diabetes especially among individuals without those comorbidities.^29,30^ The association between the DASH diet score and reduced kidney disease progression remained statistically significant even after adjusting for hypertension, suggesting additional mechanisms beyond blood pressure control contribute to the diet’s protective nature. This finding suggests that there may be additional dietary pathways that could protect and promote optimal kidney function.

The magnitude of the protective association also varied across subgroups defined by ancestry and sex. Veterans of European ancestry demonstrated a stronger association than those of African ancestry, and the interaction was statistically significant. the protective association was directionally consistent between male and female Veterans, with no statistically significant interaction by sex, although it appears females require higher DASH diet scores than males to demonstrate the protective association with CKD. The basis for these differences are unclear from the present data but could reflect unmeasured differences in dietary patterns within DASH tertiles or differences in nutrient metabolism, which both warrant further investigation.

The partial correlation analysis identified several nutrients that characterize higher DASH adherence and potentially provides insights into mechanisms of the diet’s protective nature. Potassium, magnesium, and manganese are positively associated with the DASH diet and propose several instances in which the DASH diet could benefit or support overall kidney function.^10^ Additionally, the relationship between increased sodium intake and lower protective associations of the DASH diet strengthens the significance and recommendation of lowering sodium intake in kidney disease management and prevention.^31^ These nutrient correlations are consistent with and extend previous literature on dietary mechanisms in kidney disease progression. The protective association of potassium in terms of kidney function is supported by existing literature that demonstrates the nutrient playing a substantial role in lowering the associations of inflammation and oxidative stress which contribute to kidney function decline.^32^ The beneficial associations of magnesium also supports pre-existing literature that displays the nutrient could combat phosphate toxicity by hindering the development of calcium-phosphate crystals and protects against kidney damage.^33^ Our findings correspond with existing literature that emphasizes increased sodium leads to increased fluid retention and increased blood pressure adding strain on the kidneys and can lead to kidney disease progression.^31^ These findings further emphasize the importance of a balanced nutrient intake, reinforcing dietary recommendations that prioritize potassium-rich foods while limiting sodium consumption.^34^ Accumulating evidence indicates that the adverse renal consequences of low dietary potassium intake are not mediated exclusively through changes in blood pressure, but instead involve direct effects on kidney epithelial cells.^35^ Experimental models demonstrate that potassium deficiency induces kidney injury, inflammation, and fibrosis even when blood pressure effects are dissociated, with mechanistic studies identifying potassium-sensitive signaling pathways in proximal tubule epithelial cells that directly link reduced extracellular potassium to cellular stress, metabolic remodeling, and injury.^36,37^ Our finding regarding the protective nature of manganese is a novel finding as there have been few studies that have identified the nutrient’s specific role in kidney function decline. Thus, warranting further investigation into the specific mechanism explaining this protective association.

Our study has several substantial strengths including an incredibly large sample size with a number of participants much larger than previously published findings.^38^ The MVP is an incredibly large dataset that includes diverse individuals from various backgrounds, which maintains statistical power while adjusting for multiple covariates. This robust dataset enables in-depth subgroup analysis and improves the generalizability of our findings across various populations. Also, the calculation that we utilized for the DASH score has been extensively validated which is another strength for our study design.^21,22^ However, certain limitations should be acknowledged. For example, while the MVP is incredibly informative, the cohort population is predominantly male, which limits the generalizability of our findings for women. The self-reported food frequency questionnaire data could potentially subject our findings to recall bias and inaccurate reporting; however, the MVP SFFQ has been extensively validated.^19^ Also, due to the retrospective nature of our study, our observed findings are subjected to inherent confounding; however, through substantial adjustment including social determinants of health we were able to combat this confounding.

In conclusion, our findings support the argument that strict adherence to the DASH dietary pattern is a potential strategy for limiting the risk of kidney function decline, particularly in the context of primary prevention. Our observed protective association of DASH score across various subgroups, along with the identification of key beneficial nutrients, lays the groundwork for integrating dietary interventions into clinical practice to improve kidney health. Future studies to examine potential differences in DASH adherence across ancestry are needed to inform dietary recommendations in diverse groups.

## Data Availability

The data is only available from the VA upon request and requires IRB approval.

## Acknowledgement

We are grateful to the MVP study participants for their contributions to science and all the enrolling sites listed in the Core Acknowledgement (**Supplemental Table 3**) and the MVP staff and MVP program office. A.M.H. had full access to all the data in the study and assumed responsibility for data integrity. Its contents are solely the responsibility of the authors and do not necessarily represent official views for the Million Veteran Program or the Department of Veterans Affairs or the government.

## Funding acknowledgement

The work in MVP was supported by MVP000 (MVP cores) and a CSR&D merit award #I01CX001897 titled “Genetic of Kidney Disease and Hypertension in MVP II” (PI: Adriana M. Hung). A.M.H. is supported by the US Department of Veterans Affairs Clinical Sciences R&D Service grant CX001897.

## Supplemental information

Table 1. ESRD Definition

Table 2. Covariates Definitions

Table 3. MVP Acknowledgement

